# Multiomics profiling of human plasma and CSF reveals ATN derived networks and highlights causal links in Alzheimer’s disease

**DOI:** 10.1101/2022.08.05.22278457

**Authors:** Liu Shi, Jin Xu, Rebecca Green, Asger Wretlind, Jan Homann, Noel J. Buckley, Betty M. Tijms, Stephanie J. B. Vos, Christina M. Lill, Mara ten Kate, Sebastiaan Engelborghs, Kristel Sleegers, Giovanni B. Frisoni, Anders Wallin, Alberto Lleó, Julius Pop, Pablo Martinez-Lage, Johannes Streffer, Frederik Barkhof, Henrik Zetterberg, Pieter Jelle Visser, Simon Lovestone, Lars Bertram, Alejo J. Nevado-Holgado, Petroula Proitsi, Cristina Legido-Quigley

## Abstract

**INTRODUCTION:** This study employed an integrative system and causal inference approach to explore molecular signatures in blood and CSF, the Amyloid/Tau/Neurodegeneration [AT(N)] framework, MCI conversion to AD, and genetic risk for AD.

**METHODS:** Using the EMIF-AD MBD cohort, we measured 696 proteins in cerebrospinal fluid (n=371), 4001 proteins in plasma (n=972), 611 metabolites in plasma (n=696) and genotyped data in whole-blood (7,778,465 autosomal SNPs, n=936). We investigated associations: molecular modules to AT(N), module hubs with AD Polygenic Risk scores and *APOE*4 genotypes, molecular hubs to MCI conversion and probed for causality with AD using Mendelian Randomization (MR).

**RESULTS:** AT(N) framework associated key hubs were mostly proteins and few lipids. In MR analyses, Proprotein Convertase Subtilisin/Kexin Type 7 showed weak causal associations with AD, and AD was causally associated with Reticulocalbin 2 and sphingomyelins.

**DISCUSSION:** This study reveals multi-omics networks associated with AT(N) and MCI conversion and highlights AD causal candidates.

## 1. Introduction

Alzheimer’s disease (AD) is characterised by the presence of β-amyloid (Aβ) containing plaques, and neurofibrillary tangles composed of modified tau protein together with the progressive loss of synapses and neurons [1]. The National Institute on Aging and Alzheimer’s Association (NIA-AA) have proposed to classify AD based on biomarkers of amyloid pathology (A), tau pathology (T), and neurodegeneration (N) (the ATN framework) [2]. Yet, despite their diagnostic utility, these three markers reflect only a portion of the complex pathophysiology of AD. In prodromal stages, the interplay between AT(N) changes, genetic factors and peripheral molecular changes may affect the rate of disease progression.

Conducting unbiased and high-throughput omics-based research in biological fluids and human brain tissues provides a data-driven approach to identify the many processes involved in AD pathogenesis and to prioritize links to relevant clinical and neuropathological traits. For example, an increasing number of proteomics studies [3-5], including ours [6-8], have identified AD pathophysiological pathways related to immune response and inflammation, oxidative stress, energy metabolism and mitochondrial function. Metabolomics studies have also identified such pathways related to AD [9-11]. A combination of omics, also called multi-omics or deep phenotyping studies, provides an opportunity to explore the molecular interplay with both genotypic and phenotypic variability in AD, bringing in new findings and uncovering novel pathways. Finally, causal inferences approaches allow to scrutinize the causal relationship between molecular markers and AD, highlighting potential interventional targets. Therefore, in this study, we conducted multi-omics analyses with four modalities (cerebrospinal fluid [CSF] proteomics, plasma proteomics, plasma metabolomics and whole blood genetics) from the EMIF-AD MBD study, followed by Mendelian Randomization (MR) analyses (**Figure 1**).

**Figure 1.**
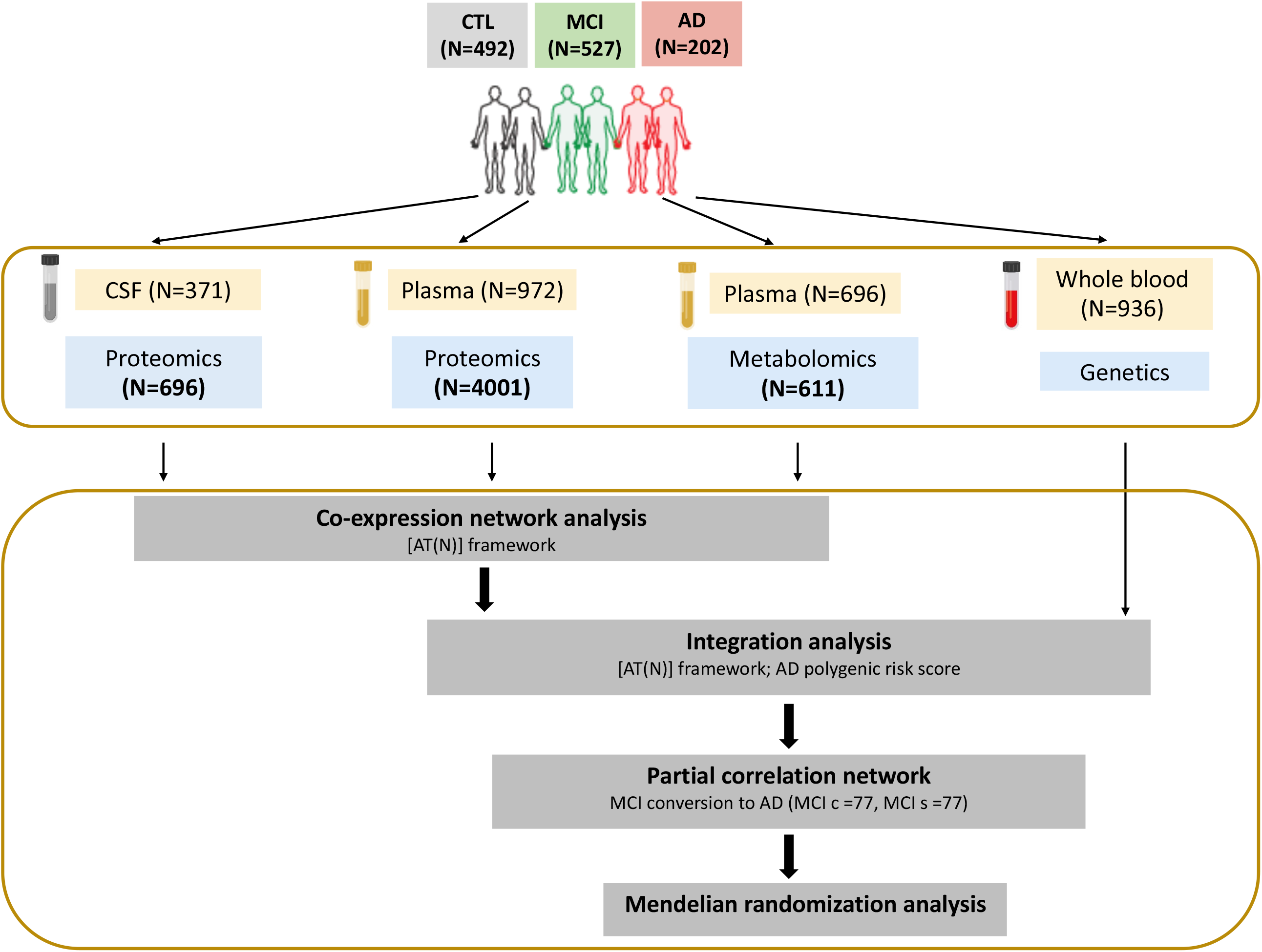
Flowchart of study design. CTL, cognitively normal controls; MCI, mild cognitive impairment; AD, Alzheimer’s disease; Aβ, β-amyloid; CSF, cerebrospinal fluid; A, amyloid pathology; T, tau pathology; N, neurodegeneration; PRS, polygenic risk score; MR, mendelian randomization.

We have four objectives: Firstly, we wanted to test if proteomic and metabolomic molecular signatures were associated with AD endophenotypes including amyloid, CSF total tau (T-tau), CSF phosphorylated tau (P-tau), white matter hyperintensity volume, CSF YKL-40, mini mental state examination (MMSE) score, and mild cognitive impairment (MCI) conversion. Secondly, we wanted to investigate the associations between molecular signatures and molecular hubs (main molecules driving associations) with *APOE*4 genotypes and AD polygenic risk scores (PRS). Thirdly, we wanted to query our findings in prodromal AD by extracting and integrating hub molecules in MCI individuals that converted to AD by computing a network for MCI converters versus non-converters. Finally, MR analyses interrogated the causal relationship between hub molecules and AD.

## 2. Methods

### 2.1. Participants: EMIF-AD Multimodal Biomarker Discovery (MBD) study

The EMIF-AD MBD study is part of the European Medical Information Framework for Alzheimer’s disease (EMIF; http://www.emif.eu/emif-ad-2/); a public-private partnership funded through the Innovative Medicines Initiative (IMI). The design of the EMIF-AD MBD study has been described previously [12]. Briefly, 1221 samples from three groups of people (cognitively normal controls [CTL], MCI and AD) were chosen from pre-existing cohorts with the goal of including samples from people with pathology as well as those without. All participating centres have agreed to share data as part of the EMIF-AD MBD study.

General clinical and demographic information were available for all subjects (including *APOE* ε4 genotype data). Furthermore, each participant had a measure of brain amyloid load, using either CSF Aβ or amyloid positron emission tomography (PET) imaging. CSF T-tau and P-tau analysis data were available for over 90% of the subjects. We used CSF (or where not available, PET) amyloid as “A”, CSF P-tau 181 as “T” and CSF T-tau as “N” to define the AT(N) framework. The classification of the status (abnormal/normal) of amyloid, P-tau and T-tau has been described previously [12]. We dichotomized these biomarkers as normal or abnormal and categorized them into four groups: no pathology (A-T-N-, referring as “A-TN-”), amyloid positive but both T and N negative (A+T-N-, referring as “A+TN-”), amyloid positive and T/N positive (including A+T-N+, A+T+N- and A+T+N+, referring as “A+TN+”) and Suspected Non-Alzheimer Pathology (SNAP, including A-T-N+, A-T+N- and A-T+N+). In addition, the following AD-related endophenotypes were also measured for the majority of the subjects: (i) CSF YKL-40; (ii) MRI measures of white matter hyperintensities; (iii) clinical assessments including baseline diagnosis, baseline MMSE score and MCI conversion [12].

### 2.2. Omics analyses

We performed multi-omics analyses for these subjects including CSF proteomics, plasma proteomics and metabolomics as well as genome-wide SNP genotyping analyses (**Figure 1**).

#### CSF proteomics

We used tandem mass tag (TMT) technique to measure proteins in CSF. More details can be found in [13]. We imputed proteins using K-nearest neighbour (K=10) and removed any missing > 70%, leading to a total of 696 proteins in 371 samples for further analysis.

#### Plasma proteomics

We used the SOMAscan assay platform (SomaLogic Inc.) to measure proteins in plasma. SOMAscan is an aptamer-based assay allowing for the simultaneous measurement and quantification of large number of proteins. Here we measured 4001 proteins in 972 individuals. The details have been described previously [14].

#### Plasma metabolomics

We measured plasma metabolites using Metabolon platform (Metabolon Inc.). Metabolites with more than 70% missing were excluded and we imputed the missing metabolites using K-nearest neighbour (K=10), resulting in 611 metabolites in 696 subjects for further analysis. More details can be found in [11].

#### Single nucleotide polymorphism (SNP) genotyping

A detailed account of the genotyping procedures and subsequent bioinformatic workflows can be found in [15]. Briefly, a total of 936 DNA samples were sent for genome-wide SNP genotyping using the Infinium Global Screening Array (GSA) with Shared Custom Content (Illumina Inc.). After quality control (QC) and imputation, a total of 7,778,465 autosomal SNPs with minor allele frequency (MAF) ≥0.01 were retained in 898 individuals of European ancestry for downstream analyses and genetic principal components (PCs) were computed [15].

### 2.3. Statistical analysis

All statistical analyses were completed using R (version 4.1.2). To compare baseline cohort characteristics across three different diagnostic groups (CTL, MCI and AD), we used one-way analysis of variance (ANOVA) and chi-square tests to compare continuous and binary variables, respectively.

#### Weighted Gene Correlation Network Analysis (WGCNA)

We used the R package WGCNA [16] to construct a weighted and unsigned co-expression network for each individual omics layer. This clustering method is based on calculating correlations between paired variables. The resulting modules or groups of co-expressed analytes were used to calculate module eigenprotein/eigenmetabolite metrics. The eigenprotein/eigenmetabolite-based connectivity (kME) value was used to represent the strength of an analyte’s correlation with the module. Analytes with high intramodular kME in the top 90th percentile within a module were considered as hub proteins/metabolites.

The correlations between eigenprotein/eigenmetabolite and AD endophenotypes were calculated using Spearman’s correlation, the *p* values were corrected with false discovery rate (FDR) and corrected *p* values are presented in a heat map. Furthermore, we used one way ANOVA test to assess pairwise difference of eigenprotein/eigenmetabolite among different AT(N) framework.

#### Pathway enrichment analysis

Protein pathway enrichment analysis was performed using WebGestalt software (http://www.webgestalt.org/). Briefly, proteins within a module were assembled into a “protein list” and all proteins measured were used as “background”. This enrichment analysis was performed on the KEGG database. Metabolite enrichment analysis was performed using the hypergeometric test. The original 60 sub-pathways pre-defined by Metabolon based on the KEGG database were employed as reference [17]. We further performed cell type enrichment analysis for CSF proteins using BEST tool (http://best.psych.ac.cn/#).

#### AD polygenic risk score (PRS) calculation

The genome-wide association study summary statistics from Kunkle et al. [18] (N=63,926; 21,982 AD clinically ascertained cases, 41,944 controls) were used as the reference data. PRS were constructed using PRSice-2 [19], with and without SNPs in the *APOE* region (chr 19, GRCh37 coordinates 44912079 to 45912079) [20]. AD PRS were computed using two *p-*value thresholds (P_T_), previously recommended for PRS including and excluding the *APOE* region: 5×10^−8^ (*APOE* region included) and 0.1 (*APOE* region excluded) [21]. SNPs in linkage disequilibrium (r2>0.001 within a 250kb window) were clumped, retaining the SNP with the lowest *p-*value.

#### Association of AD PRS and AT(N) with modules and hubs

We used linear regression analyses to investigate the association of AD PRS (as predictor) with eigenprotein/eigenmetabolite of AT(N) framework-related modules and hub proteins/metabolites (with kME varying from top 90th percentile to top 98th percentile) in these modules, adjusting for sex, age, and genetic PC1 to PC5 [22] (to control for population stratification). We used logistic regression analyses to explore the association of AT(N) markers (as binary outcome) with hubs, adjusting for sex, age and *APOE* ε4 genotype.

#### Partial correlation network

We used age, sex, *APOE* genotype, AD PRS (P_T_ = 0.1, *APOE* region excluded) and all hub proteins/metabolites (with kME in the top 90th percentile) as input features for the graphical LASSO algorithm and extended Bayesian information criterion to determine the model complexity for MCI conversion using the R package ‘huge’ [23]. Data were auto-scaled prior to model-fitting. Partial correlation network of selected metabolites, proteins and genetic variables was computed and visualized with R package ‘qgraph’.

#### Mendelian randomization

We finally investigated whether any of the A/T/N hubs correlating with MCI conversion status were causally linked to AD, by performing bi-directional two-sample Mendelian Randomization (MR) analyses implemented in the “TwoSampleMR” R package [24] and the MendelianRandomization package [25]. A number of sensitivity analyses for both single cis instrument MR and multiple (cis) instruments MR (**Supplementary methods**) were applied to determine the robustness of the MR findings.

## 3. Results

### 3.1. Subject demographics

**Table 1** shows the demographic information of subjects for each individual omics analysis. Despite the difference in sample size for each omics layer analysis, no significant difference was observed in the distribution of sex across different diagnostic groups. However, the CTL group was younger and had a lower proportion of *APOE* ε4 carriers compared with the MCI and AD groups. Furthermore, the CTL participants had longer education and higher MMSE score. In terms of AD pathology markers, the ratio of abnormality of amyloid, P-tau and T-tau in AD and MCI individuals was, as expected, significantly higher than in controls.

**Table 1.**
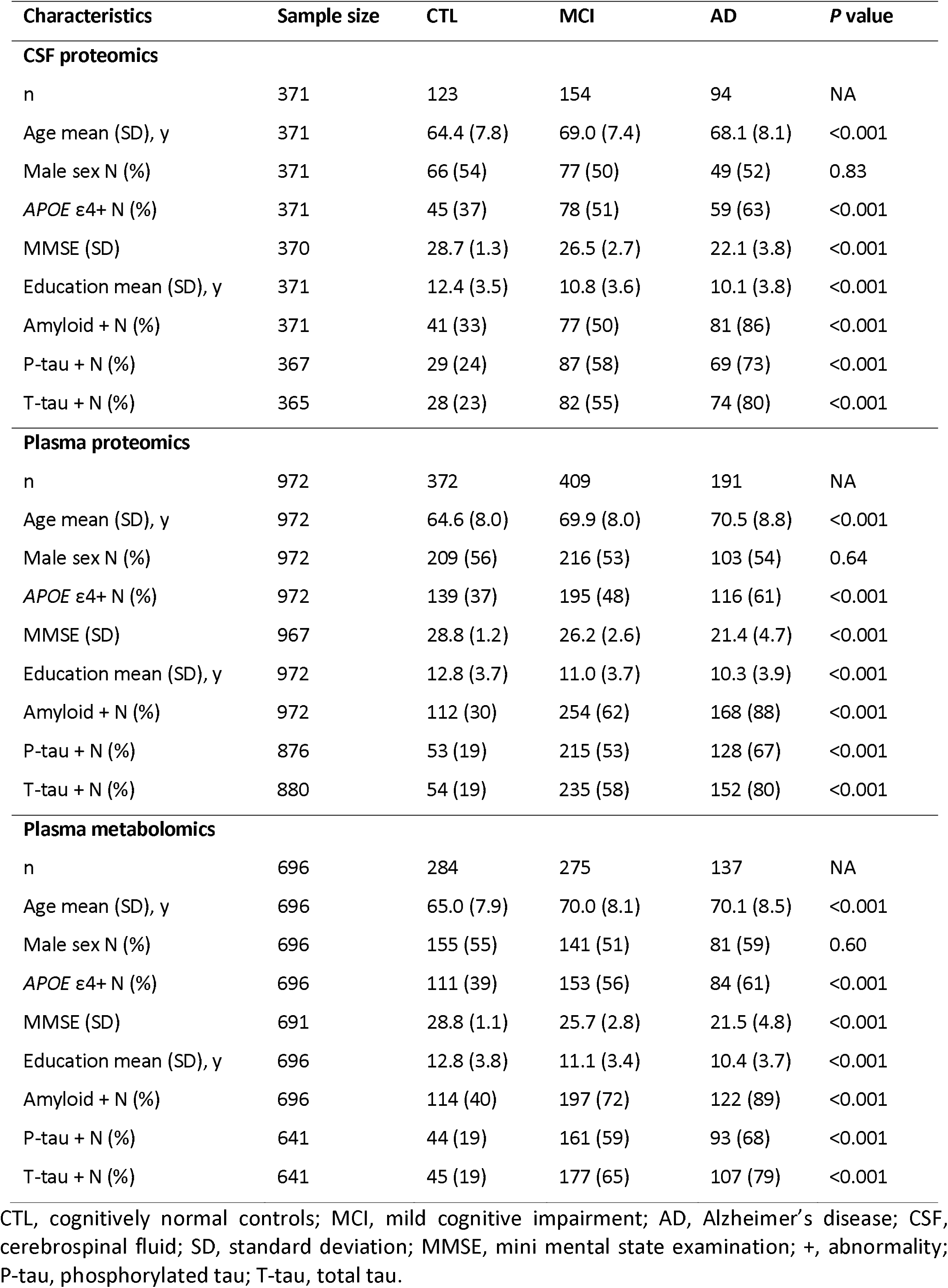
Demographics of participants included in multi-omics analysis by diagnosis. One-way analysis of variance (ANOVA) and chi-square tests were used to compare continuous and binary variables, respectively. Percentage of cases is shown in brackets for male sex, *APOE* ε4 carriers and the abnormality of amyloid, P-tau and T-tau.

| Characteristics | Sample size | CTL | MCI | AD | P value |
| --- | --- | --- | --- | --- | --- |
| **CSF proteomics** |  |  |  |  |  |
| n | 371 | 123 | 154 | 94 | NA |
| Age mean (SD), y | 371 | 64.4 (7.8) | 69.0 (7.4) | 68.1 (8.1) | <0.001 |
| Male sex N (%) | 371 | 66 (54) | 77 (50) | 49 (52) | 0.83 |
| *APOE* $\epsilon 4$+ N (%) | 371 | 45 (37) | 78 (51) | 59 (63) | <0.001 |
| MMSE (SD) | 370 | 28.7 (1.3) | 26.5 (2.7) | 22.1 (3.8) | <0.001 |
| Education mean (SD), y | 371 | 12.4 (3.5) | 10.8 (3.6) | 10.1 (3.8) | <0.001 |
| Amyloid + N (%) | 371 | 41 (33) | 77 (50) | 81 (86) | <0.001 |
| P-tau + N (%) | 367 | 29 (24) | 87 (58) | 69 (73) | <0.001 |
| T-tau + N (%) | 365 | 28 (23) | 82 (55) | 74 (80) | <0.001 |
| **Plasma proteomics** |  |  |  |  |  |
| n | 972 | 372 | 409 | 191 | NA |
| Age mean (SD), y | 972 | 64.6 (8.0) | 69.9 (8.0) | 70.5 (8.8) | <0.001 |
| Male sex N (%) | 972 | 209 (56) | 216 (53) | 103 (54) | 0.64 |
| *APOE* $\epsilon 4$+ N (%) | 972 | 139 (37) | 195 (48) | 116 (61) | <0.001 |
| MMSE (SD) | 967 | 28.8 (1.2) | 26.2 (2.6) | 21.4 (4.7) | <0.001 |
| Education mean (SD), y | 972 | 12.8 (3.7) | 11.0 (3.7) | 10.3 (3.9) | <0.001 |
| Amyloid + N (%) | 972 | 112 (30) | 254 (62) | 168 (88) | <0.001 |
| P-tau + N (%) | 876 | 53 (19) | 215 (53) | 128 (67) | <0.001 |
| T-tau + N (%) | 880 | 54 (19) | 235 (58) | 152 (80) | <0.001 |
| **Plasma metabolomics** |  |  |  |  |  |
| n | 696 | 284 | 275 | 137 | NA |
| Age mean (SD), y | 696 | 65.0 (7.9) | 70.0 (8.1) | 70.1 (8.5) | <0.001 |
| Male sex N (%) | 696 | 155 (55) | 141 (51) | 81 (59) | 0.60 |
| *APOE* $\epsilon 4$+ N (%) | 696 | 111 (39) | 153 (56) | 84 (61) | <0.001 |
| MMSE (SD) | 691 | 28.8 (1.1) | 25.7 (2.8) | 21.5 (4.8) | <0.001 |
| Education mean (SD), y | 696 | 12.8 (3.8) | 11.1 (3.4) | 10.4 (3.7) | <0.001 |
| Amyloid + N (%) | 696 | 114 (40) | 197 (72) | 122 (89) | <0.001 |
| P-tau + N (%) | 641 | 44 (19) | 161 (59) | 93 (68) | <0.001 |
| T-tau + N (%) | 641 | 45 (19) | 177 (65) | 107 (79) | <0.001 |

CTL, cognitively normal controls; MCI, mild cognitive impairment; AD, Alzheimer's disease; CSF, cerebrospinal fluid; SD, standard deviation; MMSE, mini mental state examination; +, abnormality; P-tau, phosphorylated tau; T-tau, total tau.

### 3.2 Co-expression network analysis of individual omics modalities reveals modules linked to AD endophenotypes

We first performed a clustering analysis of the CSF proteome using WGCNA. We found four modules (M) of co-expressed proteins. We ranked modules based on size from largest (M1 turquoise; n= 526 proteins) to smallest (M4 yellow; n=51 proteins) (**Figure 2A**, Table S1). We further investigated the biological significance of proteins in each module and found that three modules (M1 turquoise, M2 blue and M4 yellow modules) were enriched with various pathways after FDR correction (**Figure 2B**). When checking cell type enrichment, we found that all four modules were enriched with endothelial cells. Furthermore, M1 turquoise module was enriched with oligodendrocytes, neurons and astrocytes. M2 blue and M4 yellow modules were enriched with microglia (**Figure 2A**).

**Figure 2.**
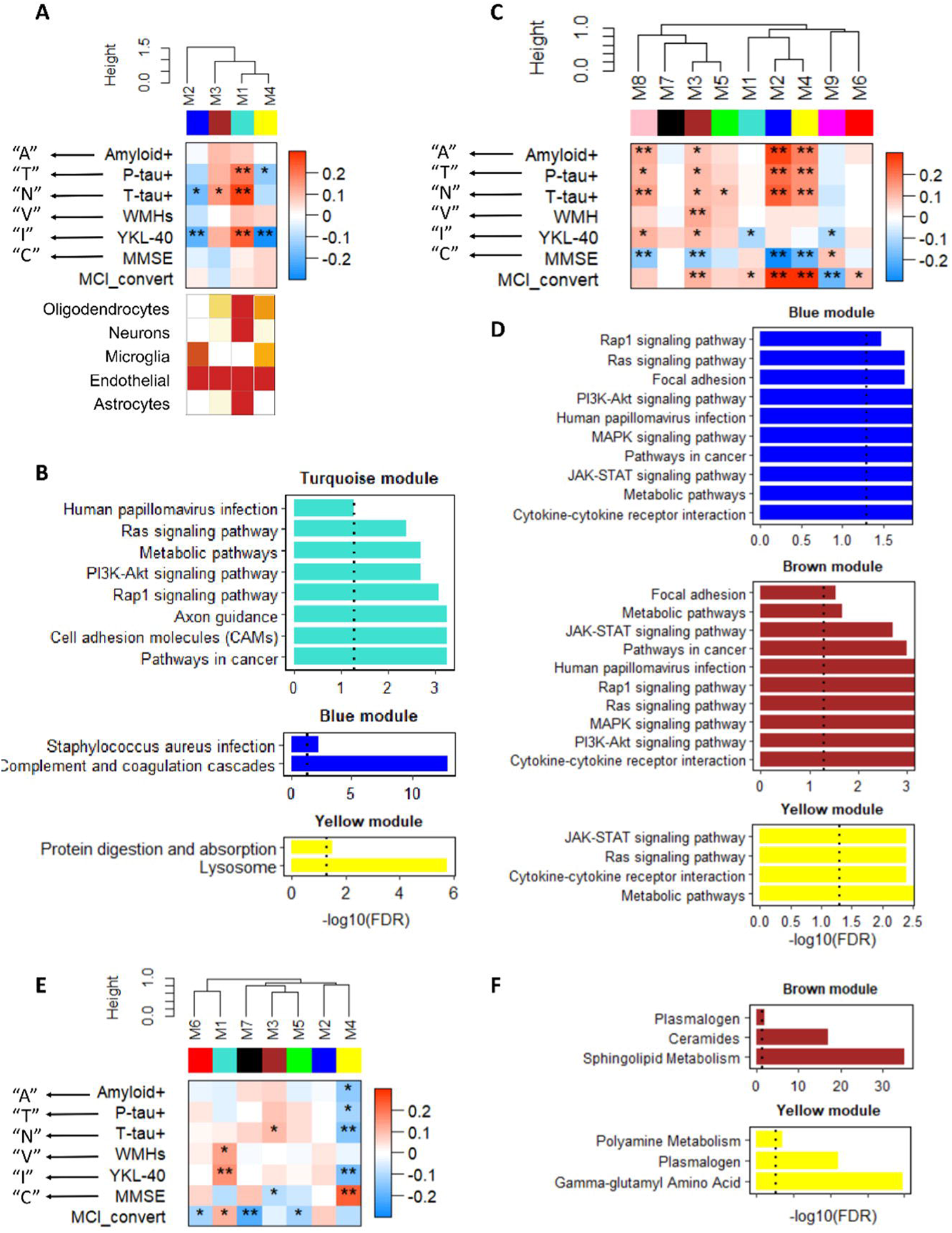
Individual omics modules correlating to AD endophenotypes. (A) Weighted gene correlation network analysis (WGCNA) of the CSF proteomics and cell type enrichment analysis of modules; (B) Enriched KEGG pathways of three modules in CSF proteins; (C) WGCNA of plasma proteomics; (D) Enriched KEGG pathways of three modules in plasma proteins; (E) WGCNA of plasma metabolomics; (F) Enriched KEGG pathways of two modules in plasma metabolites. * and ** denote significant correlations *p* < 0.05 and *p* < 0.001 after false discovery rate (FDR) correction respectively; CSF, cerebrospinal fluid; “A”, amyloid; “T”, tau; “N”, neurodegeneration; “V”, vascular; “I”, inflammation; “C”, cognition; +, abnormality; P-tau, phosphorylated tau; T-tau, total tau; WMH, white matter hyperintensity; MMSE, mini mental state examination; MCI, mild cognitive impairment.

We then assessed the module correlations to AD endophenotypes. We used amyloid-β as “A”, CSF P-tau levels as a biomarker of tau (“T”), CSF T-tau as biomarkers of neurodegeneration (“N”), white matter hyperintensity (WMH) volume as a biomarker for vascular disease burden (“V”), CSF YKL-40 as a biomarker of inflammation (“I”) and MMSE score as “C” (**Figure 2A**). Overall, two (M1 and M4) and three (M1, M2 and M3) modules were significantly associated with “T” and “N”, respectively after FDR correction. Furthermore, three (M1, M2 and M4) modules were associated with “I”. None of the modules were correlated with “A”, “V”, “C” or MCI conversion.

We used the same approach to analyse plasma proteomics and metabolomics data. We obtained nine modules from plasma proteins (**Figure 2C**, previously published [26]). Four modules (M2, M3, M4 and M8) had positive correlations with “A”, “T” and “N”. One (M3) and four (M1, M3, M8 and M9) modules were associated with “V” and “I”, respectively. In comparison, most plasma modules were associated with “C” and MCI conversion. Furthermore, such associations were in concordance with AT(N) markers correlations. For example, M2, M3, M4 and M8 modules were positively associated with “A”, “T” and “N” but were negatively correlated with MMSE score. Furthermore, they were increased in MCI converters (n=103) compared with MCI non-converters (n=223) (**Figure 2C**). We further investigated the biological significance of proteins in four AT(N) markers-related modules (M2, M3, M4 and M8) and found that three of them were enriched with various pathways, such as cytokine-cytokine receptor interaction and metabolic pathways (**Figure 2D**).

For plasma metabolomics, we obtained seven modules (**Figure 2E**), among which M4 module was negatively associated with “A”, “T” and “N” and M3 module was positively associated with “N”. Furthermore, one (M1) and two (M1 and M4) modules were associated with “V” and “I”, respectively. Two (M3 and M4) and four (M1, M5, M6 and M7) modules were associated with “C” and MCI conversion respectively. Furthermore, such associations were in concordance with AT(N) markers correlations. We further investigated the biological significance of metabolites in AT(N) markers-related modules (M3 and M4) and found that they were enriched in lipid pathways (**Figure 2F**).

### 3.3 Correlation of individual omics modules with the AT(N) framework

We dichotomized AT(N) biomarkers as normal or abnormal and categorized individuals into one of four groups: A-T-N-(no pathology), A+TN-(amyloid pathology), A+TN+ (Alzheimer pathology) and A-TN+ (SNAP). We then assessed the expression of each module eigenprotein/eigenmetabolite across different ATN groups. For CSF protein modules, we found that three modules (M1 turquoise, M2 blue and M4 yellow) showed a significant difference across ATN profiles from one-way ANOVA test (**Figure 3A-C**). Four plasma protein modules (M2 blue, M3 brown, M4 yellow and M8 pink, **Figure 3D-G**, adapted from [26]) and three plasma metabolites modules (M4 yellow, M5 green and M3 brown, **Figure 3H-J**) showed a significant difference across ATN profiles.

**Figure 3.**
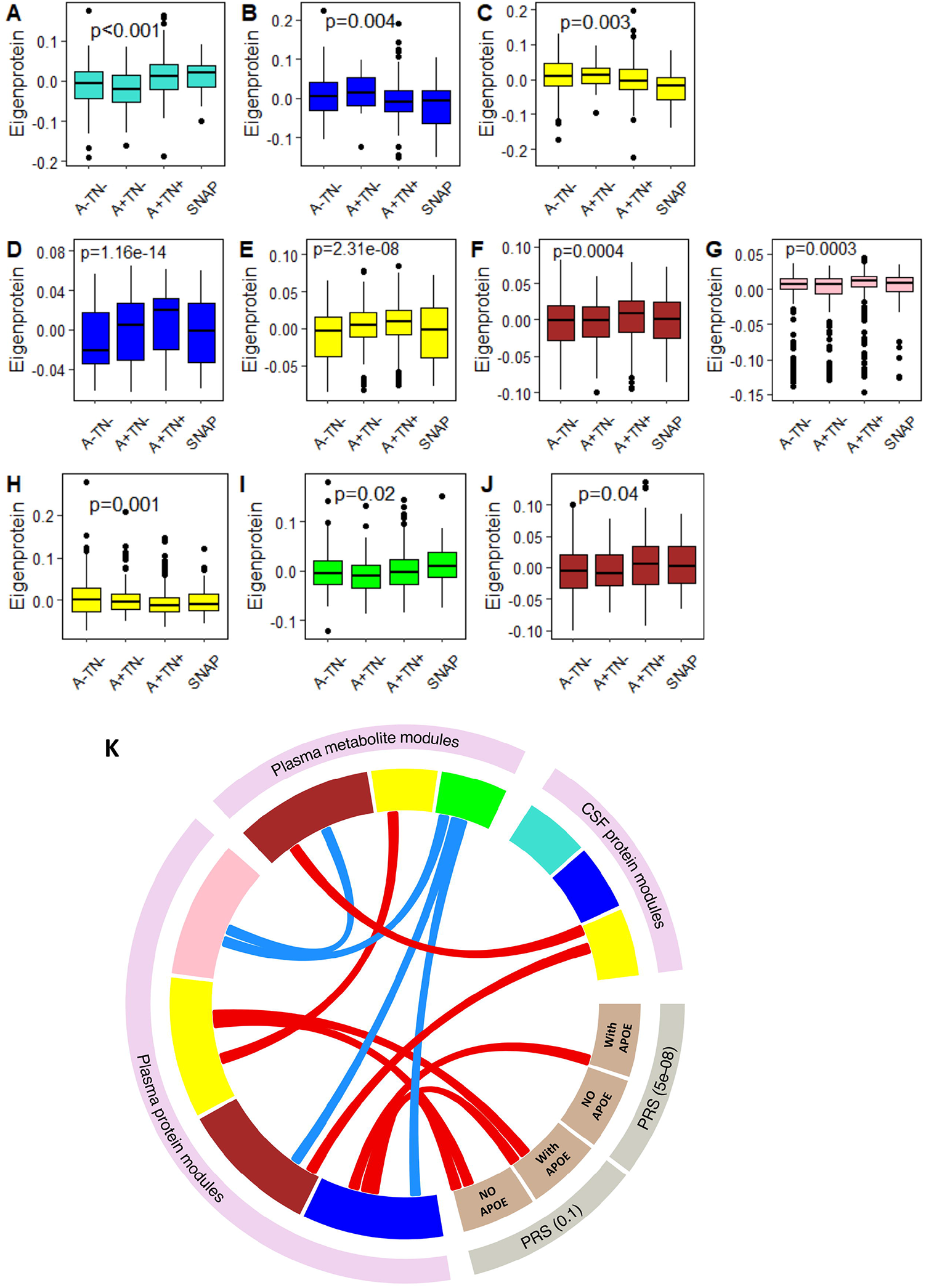
Protein and metabolite modules correlate to AT(N) profile and AD PRS. The relationship of the AT(N) framework with (A-C) three CSF protein modules, (D-G) four plasma protein modules, (H-J) three plasma metabolite modules, (K) Relation of AT(N) framework-related modules with AD PRS (with and without *APOE* region) at two thresholds (PT=5×10^−8^ & 0.1); red and blue links denoted positive and negative correlations, respectively. CSF, cerebrospinal fluid; SNAP, Suspected Non-Alzheimer Pathology.

### 3.4 Association between AT(N) framework-related modules and AD PRS

We firstly selected AT(N) framework-related modules from each individual omics for further analysis. As a result, we selected three CSF protein modules (M1 turquoise, M2 blue and M4 yellow), four plasma protein modules (M2 blue, M3 brown, M4 yellow and M8 pink) and three plasma metabolite modules (M3 brown, M4 yellow and M5 green). We then analysed the correlations between these ten modules as well as between these modules and AD PRS. When analysing associations between modules, we found that the metabolite M5 green module was negatively correlated with three plasma protein modules (M2 blue, M3 brown and M8 pink). Additionally, a negative correlation was observed between metabolite M3 brown module and plasma protein M8 pink module. In contrast, a positive correlation was observed between metabolite M4 yellow module and plasma protein M4 yellow module as well as between metabolite M3 brown module and CSF protein M4 yellow module. In addition, CSF protein M4 yellow module was positively associated with plasma protein M3 brown module (**Figure 3K, Table S1-S3**).

When analysing the associations between these modules and AD PRS, we found that only plasma protein modules were significantly associated with AD PRS. In detail, two plasma protein modules (M2 blue and M4 yellow) were positively associated with PRS (*APOE* region included and excluded) at P_T_ =0.1. Additionally, M2 blue module was significantly associated with PRS at 5×10^−8^ threshold with *APOE* gene region included (**Figure 3K, Table S4**).

### 3.5 Association of hub proteins/metabolites with AT(N) markers and AD PRS

We selected hub proteins/metabolites within AT(N) framework-related modules and analysed the association between these hub proteins/metabolites (with kME varying from top 90th percentile to top 98th percentile, **Table S5**), as well as the association of these hub proteins/metabolite with AT(N) markers and AD PRS. When checking the associations between hub metabolites and proteins, we found that there was a strong correlation between metabolites and plasma proteins. In detail, five metabolites (four phosphatidylethanolamines (PEs) and one LysoPE) correlated with most hub proteins after controlling for multiple testing. Two metabolites (sphingomyelins [SM] d40:2 and d41:2) in M3 brown module were correlated with proteins in plasma M8 pink module and CSF M4 yellow module. In contrast, relatively week correlations were observed between CSF and plasma proteins. (**Figure 4A, Table S6**).

**Figure 4.**
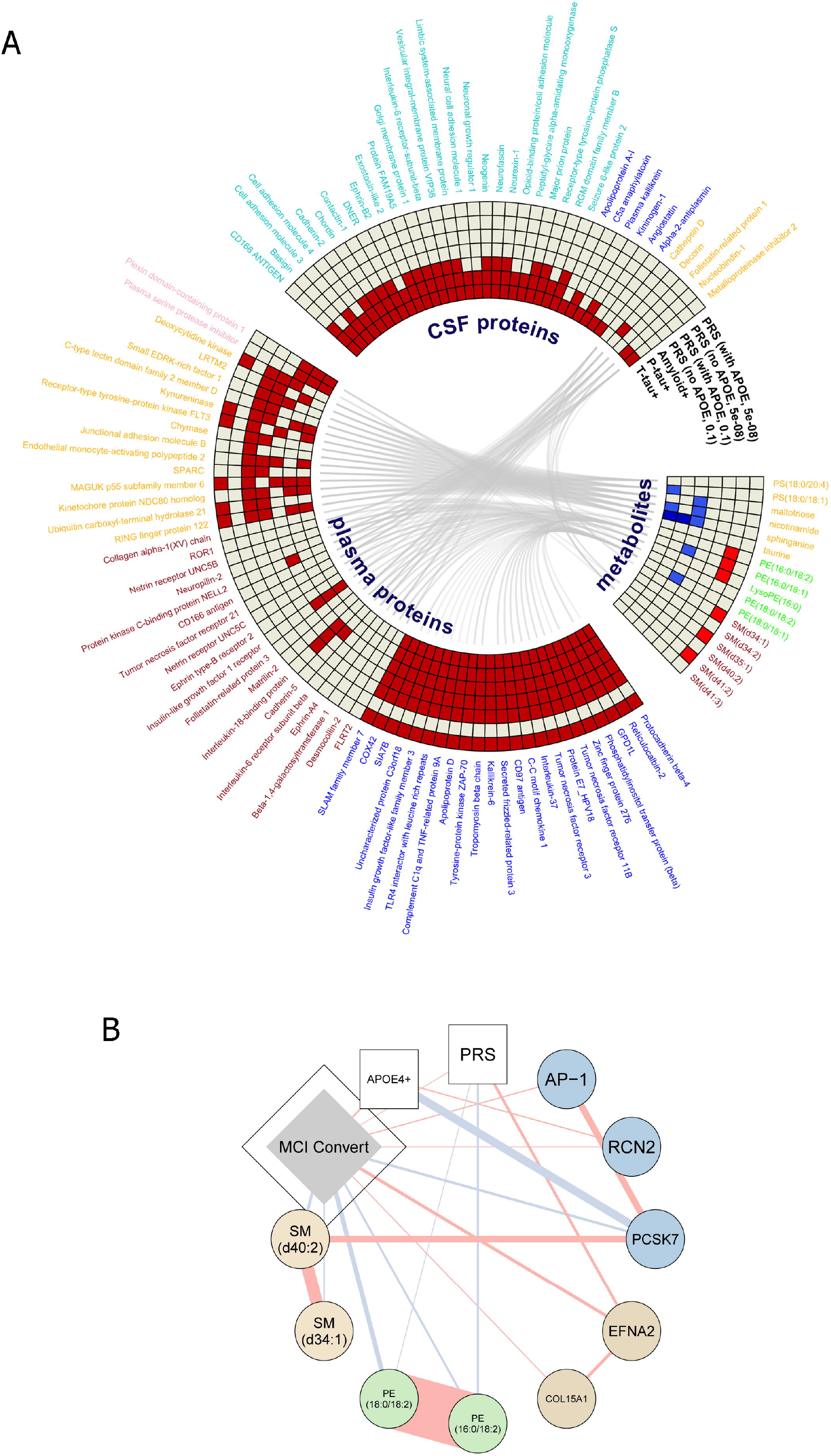
(A) Relation of hub proteins/metabolites with AT(N) markers and PRS; hub proteins/metabolites from three CSF protein modules (M1 turquoise, M2 blue and M4 yellow), four plasma protein modules (M2 blue, M3 brown, M4 yellow and M8 pink), and three plasma metabolite modules (M4 yellow, M5 green and M3 brown); red, blue, light red and light blue squares denoted positive association at FDR level (pFDR<0.05), negative association at FDR level (pFDR<0.05), positive association at nominal level (p<0.05, pFDR>0.05), and negative association at nominal level (p<0.05, pFDR>0.05), respectively. **(B) Partial correlation network selected for hub metabolites/proteins, genetic factors, and MCI conversion**. PRS, polygenic risk score; MCI, mild cognitive impairment.

We also investigated the association of these proteins/metabolites with AD PRS (*APOE* region included and excluded (**Table S7**)). For plasma hub proteins, all 23 proteins in M2 blue module were positively associated with AD PRS both at P_T_ =5×10^−8^ (*APOE* region included) and P_T_ =0.1 (*APOE* region included and excluded). Similar trend was also observed for most proteins in M4 yellow module, with only six proteins being positively associated with AD PRS at P_T_ =5×10^−8^ (*APOE* region included), whereas most proteins, except for three, were associated with the P_T_=0.1 AD PRS (with *APOE* and without *APOE*) (**Figure 4A**). For hub metabolites, three SMs in M3 brown module and three PEs in M5 green module were associated with AD PRS (P_T_ =5×10^−8^) with and without *APOE* region respectively. However, such associations did not pass FDR correction (**Figure 4A** in light red). No associations were observed between CSF hub proteins and AD PRS.

When investigating the association of hub proteins and metabolites with AT(N) markers, we found that most CSF and plasma hub proteins were positively associated with amyloid, P-tau and T-tau after FDR correction. In contrast, hub metabolites were negatively associated with amyloid, P-tau and T-tau only at nominal level except for sphinganine (**Figure 4A**) (**Table S8**).

### 3.6 Hub molecules integration in MCI conversion

Having demonstrated the association of hub proteins/metabolites with AT(N) markers and AD PRS, we then sought to find a multimodal signal that might shed insights on MCI conversion. To do this we first used LASSO algorithm and extended Bayesian information criterion to select features from age, sex, AD PRS (P_T_ = 0.1, *APOE* region excluded), *APOE* ε4 genotype and all plasma hub metabolites/proteins (with kME in the top 90th percentile, **Table S9**) to predict MCI conversion. As a result, AD PRS, *APOE* ε4 genotype and several metabolites/proteins were selected from LASSO. Of the metabolites/proteins, two SMs, two PEs and one protein (proprotein convertase subtilisin/kexin type 7 [PCSK7]) from the blue module were negatively correlated with MCI conversion while the rest four selected proteins were positively associated with MCI conversion including reticulocalbin 2 (RCN2) from the blue module, and three proteins from the brown module: ephrin receptor tyrosine kinase A2 (EFNA2), Collagen alpha-1(XV) chain (COL15A1) and AP-1 complex subunit gamma-like 2 (AP-1) (**Figure 4B**). In addition, correlations were also observed between metabolites/proteins and AD PRS and *APOE* ε4 genotype (**Figure 4B**).

### 3.7 Causal links of hub proteins/metabolites with AD

We finally used a bidirectional two-sample Mendelian randomization to determine whether there was evidence for a causal relationship of MCI conversion related hub proteins/metabolites with Alzheimer’s disease. Using Wald ratio estimate, we observed weak associations between PCSK7 and AD as well as between COL15A1 and AD using data from Sun et al. [27]. In sensitivity analyses, the causal relationship between PCSK7 and AD was replicated using summary data from an independent protein GWA study by Suhre et al. [28] (**Table 2**). Further support for causal effects for the association of PCSK7 with AD came from multiple-cis instrument MR (p<0.001 for IVW, 95% CI = 0.8 to 0.9, N _SNPs_ = 4, **Figure S4**), although this was not the case for COL15A1 (**Table S10**). Multiple-cis instrument MR robust methods (MR-Egger and Weighted-median MR) and sensitivity analyses estimates for PCSK7 were consistent with Wald ratio estimates in direction and magnitude, and showed no horizontal pleiotropy or evidence of heterogeneity, further supporting the validity of the MR assumptions (**Table S10**). In reverse MR analysis, we identified a causal association between Alzheimer’s disease, RCN2 and SM (**Table 2, Figure S1-3**). Robust methods and sensitivity analyses provided additional support for such causal effects (**Table S10**).

**Table 2.** Examination of the causal relationship between hub proteins/metabolites and Alzheimer’s using bidirectional Mendelian randomization.

| Protein | Estimate* |  |  |  |  |  |
| --- | --- | --- | --- | --- | --- | --- |
|  | Forward (hub proteins/metabolites<br>→Alzheimer's disease) |  |  | Backward (Alzheimer's disease→ hub<br>proteins/metabolites) |  |  |
|  | No. of SNPs | Slope (95% CI) | P value | No. of SNPs | Slope (95% CI) | P value |
| PCSK7 | 1 [27] | 0.88 (0.79 - 0.99) | **0.029** | 20 | 0.06 (-0.14 - 0.02) | 0.135 |
| PCSK7 | 1 [28] | 0.96 (0.93 - 0.99) | **0.027** | NA | NA | NA |
| RCN2 | 1 [27] | 1.04 (0.87 - 1.24) | 0.648 | 20 | 0.10 (0.03 - 0.17) | **0.004** |
| EFNA2 | 1 [27] | 1.04 (0.81 - 1.34) | 0.741 | 20 | -0.02 (-0.09 - 0.06) | 0.679 |
| AP-1 | 1 [27] | 0.84 (0.67 - 1.04) | 0.114 | 20 | -0.01 (-0.09 - 0.07) | 0.804 |
| COL15A1 | 1 [27] | 1.15 (1.01 - 1.30) | **0.032** | 20 | 0.02 (-0.05 - 0.09) | 0.544 |
| SM | 58 [29] | 1.28 (0.87 - 1.89) | 0.212** | 21 | 0.14 (0.03 - 0.24) *** | **0.012** |

\*The Wald ratio estimate and inverse variance weighting (IVW) estimate were used for MR analyses with a single and multiple SNPs, respectively.
\*\* Cochran's Q $p < 0.001$
\*\*\*there was significant heterogeneity even after removing APOE (Cochran's Q $p < 0.05$ ).

## 4. Discussion (words:1161)

Alzheimer’s disease is characterized by non-linear and heterogeneous biological alterations. Multi-level biological networks underlie AD pathophysiology, including but not limited to proteostasis (amyloid-β and tau), synaptic homeostasis, inflammatory and immune responses, lipid and energy metabolism, and oxidative stress [30]. Therefore, a systems-level approach is needed to fully capture AD multifaceted pathophysiology. Here we used unbiased and high throughput multi-omics profiling of AD. We applied correlation network analysis to identify modules linked to a variety of AD endophenotypes including “A”, “T”, “N”, “V”, “I” and “C”. We found that four modules obtained from CSF proteins were associated with at least one pathology marker of “T” (P-tau), “N” (T-tau) and “I” (YKL-40). Furthermore, the three “I” related modules (M1 turquoise, M2 blue and M4 yellow) were enriched with either microglia or astrocytes, which are key cellular drivers and regulators of neuroinflammation [31], further indicating the consistency between correlation network analysis and cell type enrichment analysis. In addition, of the four modules, three were enriched with various pathways which have been reported being associated with Alzheimer’s such as Ras signalling pathway [32], axon guidance [33], cell adhesion molecules (CAMs) [34], and lysosome pathway [35], further demonstrating the relatedness of these proteins with AD.

From plasma metabolomics, we found that the M3 brown module was associated with “N” (T-tau) and “C” (cognition) and enriched with sphingolipid and ceramide metabolism. These findings align with literature report as the lipids within this module have been reported being associated with cognitive progression [36] and hippocampal atrophy [37]. In addition, M4 yellow module was associated with five AD pathology markers (“A”, “T”, “N”, “I” and “C”) and enriched with three pathways including gamma-glutamyl amino acid, plasmalogen, and polyamine metabolism. The findings are also consistent with previous reports showing that these pathways were associated with AD pathogenesis [38] and inflammatory cascade [39].

Two modules (M2 blue and M4 yellow) from plasma proteomics were associated with AD PRS (both with and without *APOE* gene) at 0.1 level. Of the two modules, M2 blue module was associated with PRS at 5×10^−8^ thresholds only when PRS included SNPs in the *APOE* region, indicating that such association may be driven by *APOE*. Hub proteins in M2 blue module were correlated with PRS at 5×10^−8^ thresholds only with SNPs in the *APOE* region included, further indicating that associations may be driven by *APOE*. For plasma metabolomics, three sphingomyelins (SMs) from M3 brown module were associated PRS (P_T_ = 5×10^−8^) nominally only when the *APOE* region was included, also indicating *APOE* gene dependence. This is in line with literature findings that nominal association between SMs and PRS was reported [40].

We identified several closely correlated networks for metabolites, proteins, genetic factors, and MCI conversion. Interestingly *APOE* and MCI conversion status were correlated to PCSK7 and sphingomyelins SMs have been previously associated with cognitive progression in AD [41-43]. Furthermore, the integration of AD PRS showed that phosphatidylethanolamines [44, 45] and EFNA2 [46] were associated to both (MCI converter and AD PRS), with potential as early targets.

We finally investigated the causal relationship between A/T/N hubs associated with MCI conversion status and AD. Our MR analyses highlighted a potential weak causal relationship between PCSK7 and AD which was robust in both single and multiple cis instruments MR analyses and was replicated using an independent pQTL dataset. We also found a causal relationship in the opposite direction, whereby AD status is potentially causally linked to RCN2 that has been proposed as a therapeutic target for atherosclerosis [47]. Finally, although we didn’t have GWA summary data for the SM and PE hubs examined in this study, our MR analyses showed that AD was causally linked to SM levels, as previously shown when NMR data were used [48].

These findings are of great translational potential, particularly PCSK7 for which studies in Alzheimer’s disease are lacking. This convertase protein is very interesting as it is found in the BACE1 locus region which encompasses several genes (PCSK7, RNF214, BACE1, CEP164) making it a plausible protein activator of downstream amyloid deposition [49].

The causal associations from AD to RCN2 and sphingomyelins are also intriguing as both highlight a possible vascular component caused by AD genetic liability, bringing new directionality between vascular disease and dementia. These molecules and their potential causal links to AD suggest novel avenues of research and intervention.

There are limitations for our study. First, the population in this study is of European ancestry and mainly included participants who had high ratio of amyloid pathology and *APOE* ε4 carriers. Therefore, they are not necessarily representative of the broader community. Validation in independent cohorts and particularly in other ethnic groups and community-based populations are needed to see if the results are generalizable.

Despite this, our study is the largest study we are aware of to report multi-omics relating to AD endophenotypes, particularly to the AT(N) framework. Our findings offer new insights into changes in individual proteins/metabolites linked to AD endophenotypes, the AT(N) framework and AD PRS. The nominated causal proteins/metabolites may be tractable targets for mechanistic studies of AD pathology. Furthermore, they may represent promising drug targets in the early stages of AD.

## Data Availability

The datasets generated and analysed during the current study are available from the EMIF-AD Catalogue via submitted research proposals which have to be approved by the data-owners from each parent cohort.

## Acknowledgements

This research was conducted as part of the EMIF-AD MBD project which has received support from the Innovative Medicines Initiative Joint Undertaking under EMIF grant agreement no 115372, resources of which are composed of financial contribution from the European Union’s Seventh Framework Programme (FP7/2007-2013) and EFPIA companies’ in-kind contribution. The DESCRIPA study was funded by the European Commission within the 5th framework program (QLRT-2001-2455). The EDAR study was funded by the European Commission within the 5th framework program (contract # 37670). The Leuven cohort was funded by the Stichting voor Alzheimer Onderzoek (grant numbers #11020, #13007 and #15005). RV is a senior clinical investigator of the Flemish Research Foundation (FWO). The San Sebastian GAP study is partially funded by the Department of Health of the Basque Government (allocation 17.0.1.08.12.0000.2.454.01.41142.001.H). We acknowledge the contribution of the personnel of the Genomic Service Facility at the VIB-U Antwerp Center for Molecular Neurology. The research at VIB-CMN is funded in part by the University of Antwerp Research Fund. HZ is a Wallenberg Scholar supported by grants from the Swedish Research Council (#2018-02532), the European Research Council (#681712), Swedish State Support for Clinical Research (#ALFGBG-720931), the Alzheimer Drug Discovery Foundation (ADDF), USA (#201809-2016862), and the UK Dementia Research Institute at UCL. FB is supported by the NIHR biomedical research centre at UCLH. LS is funded by the Virtual Brain Cloud from European commission (grant no. H2020-SC1-DTH-2018-1). R.G. was supported by the National Institute for Health Research (NIHR) Biomedical Research Centre at South London and Maudsley NHS Foundation Trust and King’s College London. This paper represents independent research part-funded by the National Institute for Health Research (NIHR) Biomedical Research Centre at South London and Maudsley NHS Foundation Trust and King’s College London. The views expressed are those of the author(s) and not necessarily those of the NHS, the NIHR or the Department of Health and Social Care. JX and CLQ thank Lundbeck Fonden for the support (grant no. R344-2020-989).

## Ethics Statement

Written informed consent was obtained from all participants before inclusion in the study. The medical ethics committee at each site approved the study.

