## Supplementary figures for "Multiomics profiling of human plasma and CSF reveals ATN derived networks and highlights causal links in Alzheimer’s disease"

### Slide 1
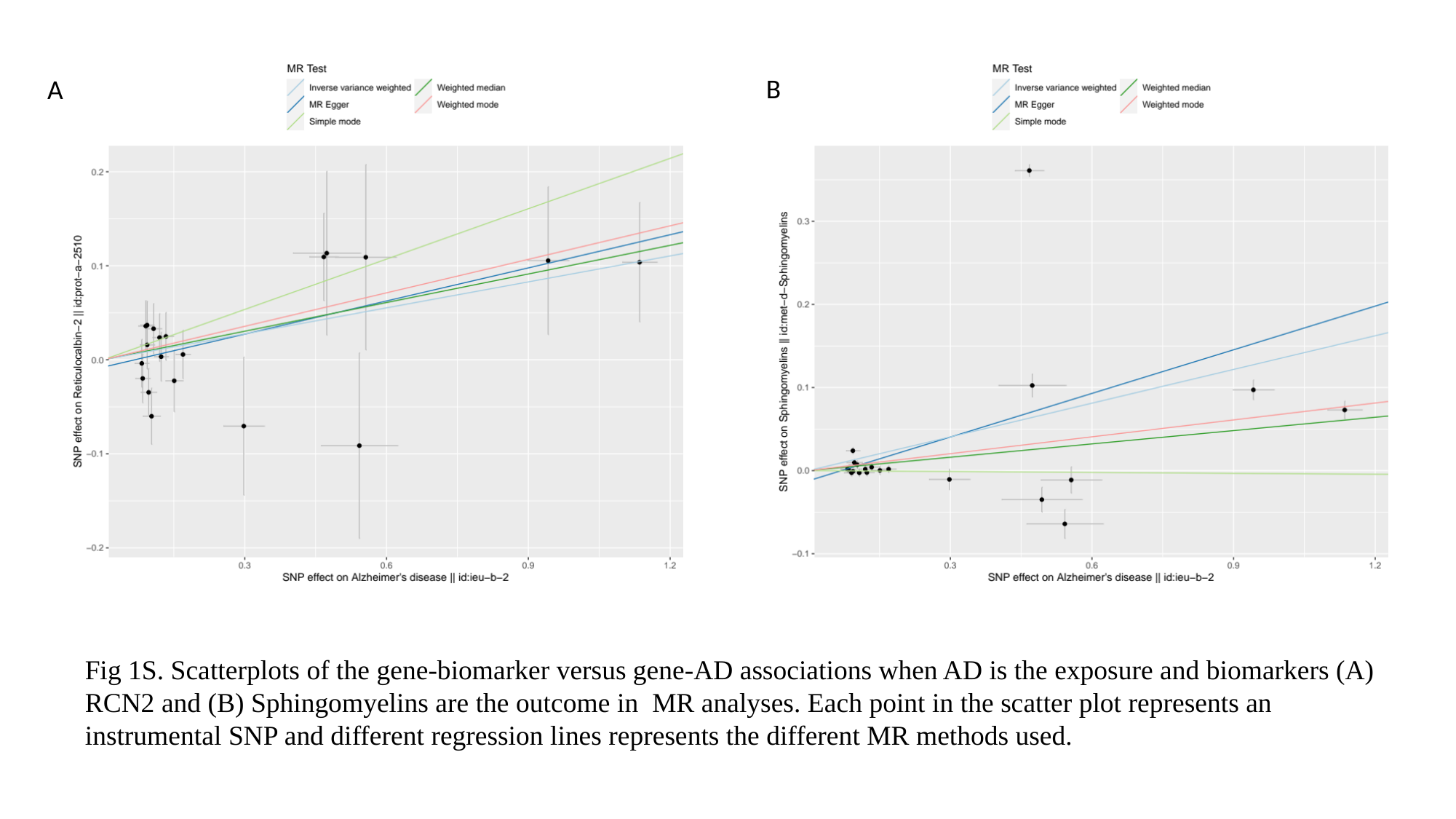

B
A
Fig 1S. Scatterplots of the gene-biomarker versus gene-AD associations when AD is the exposure and biomarkers (A) RCN2 and (B) Sphingomyelins are the outcome in  MR analyses. Each point in the scatter plot represents an instrumental SNP and different regression lines represents the different MR methods used.

### Slide 2
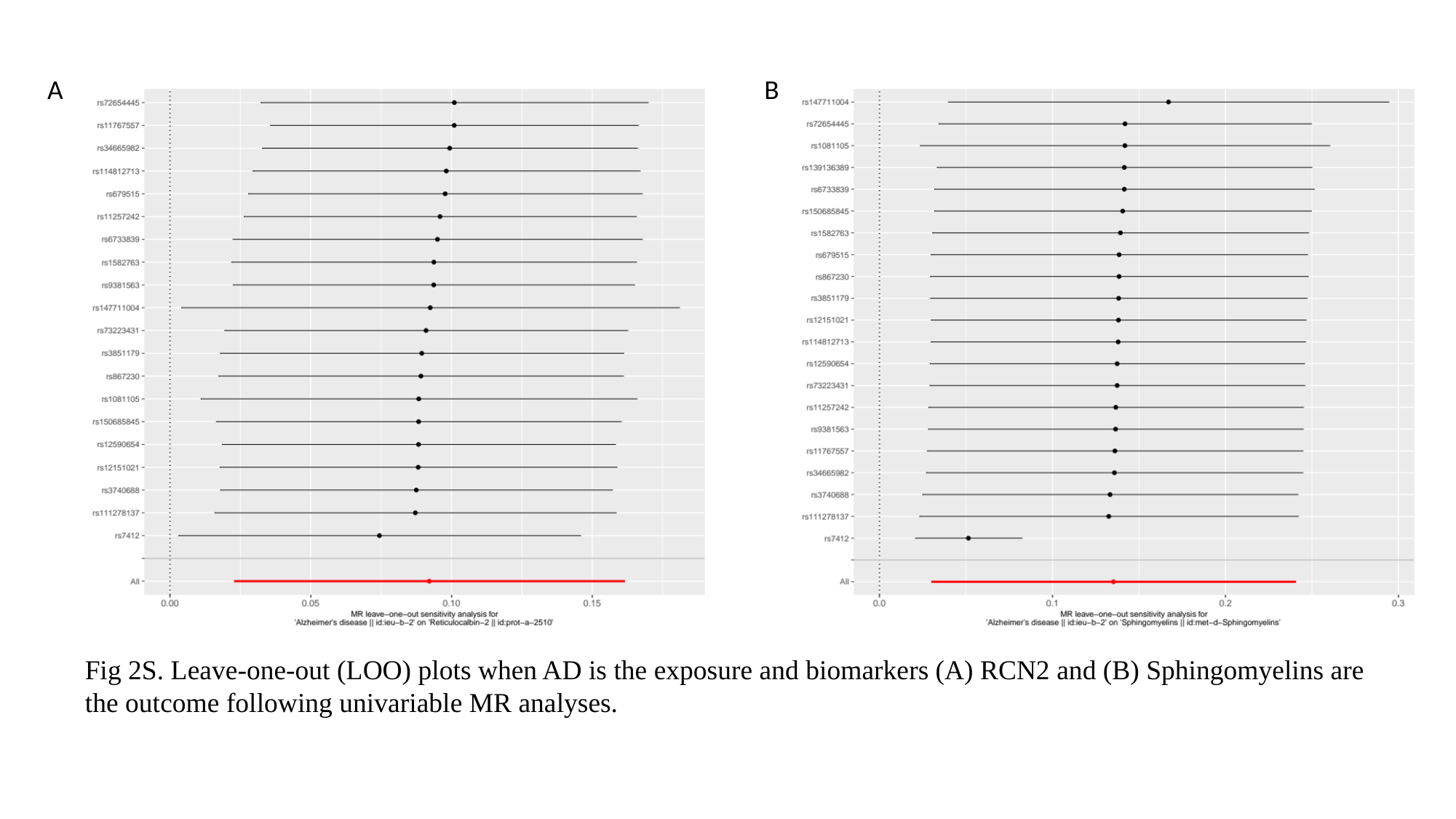

A
B
Fig 2S. Leave-one-out (LOO) plots when AD is the exposure and biomarkers (A) RCN2 and (B) Sphingomyelins are the outcome following univariable MR analyses.

### Slide 3
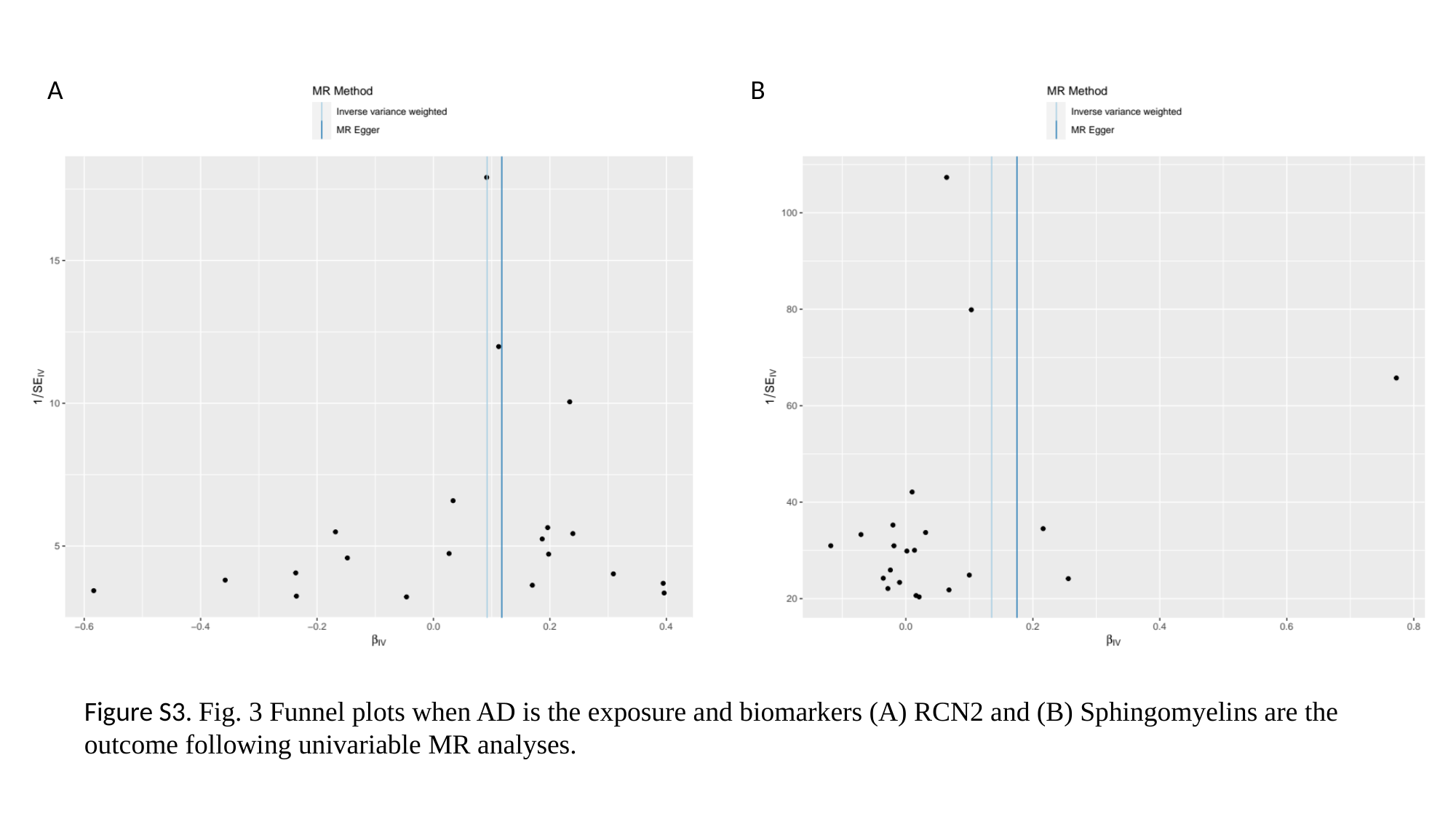

A
B
Figure S3. Fig. 3 Funnel plots when AD is the exposure and biomarkers (A) RCN2 and (B) Sphingomyelins are the outcome following univariable MR analyses.

### Slide 4
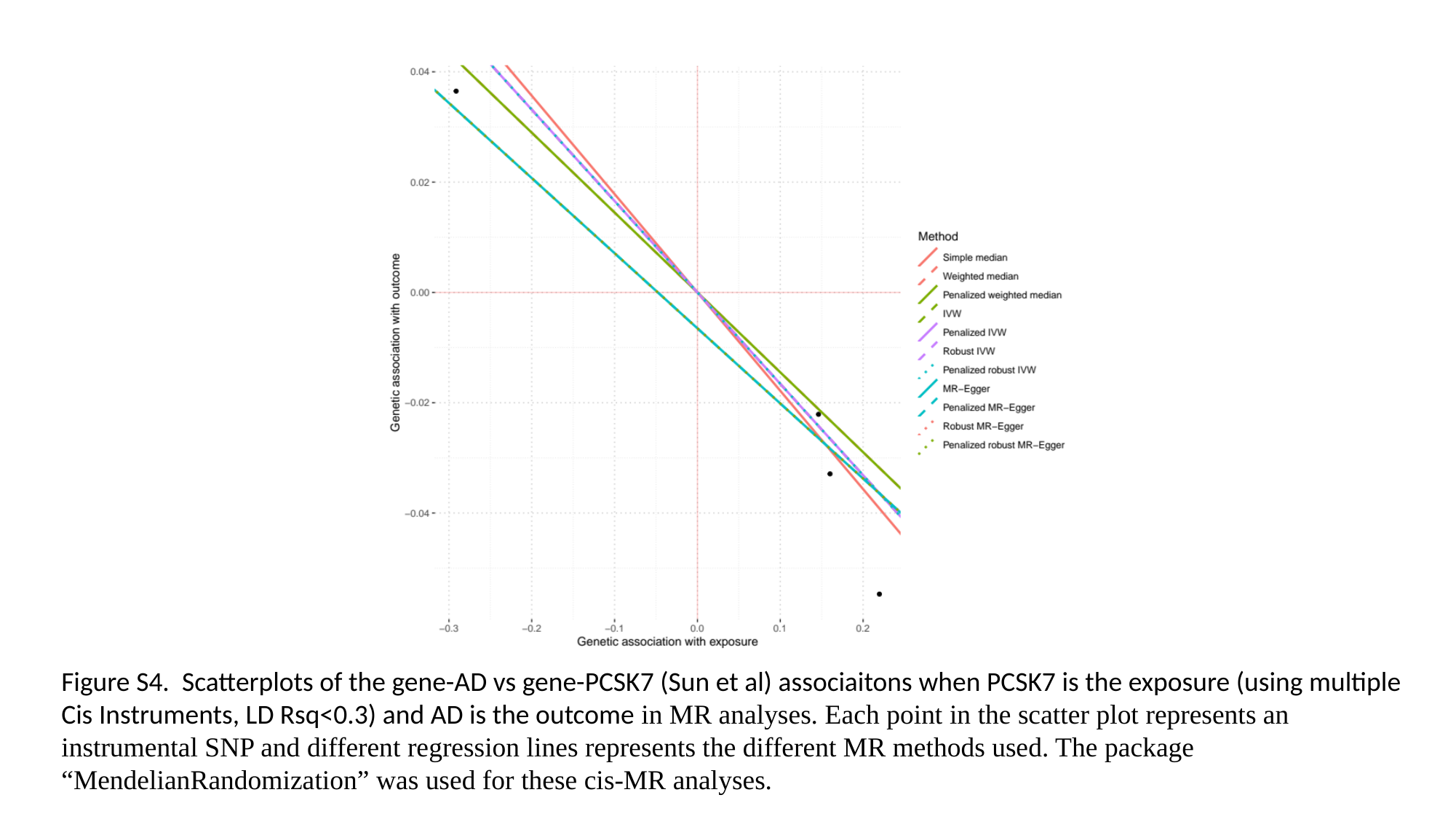

Figure S4. Scatterplots of the gene-AD vs gene-PCSK7 (Sun et al) associaitons when PCSK7 is the exposure (using multiple Cis Instruments, LD Rsq<0.3) and AD is the outcome in MR analyses. Each point in the scatter plot represents an instrumental SNP and different regression lines represents the different MR methods used. The package “MendelianRandomization” was used for these cis-MR analyses.
