## Supplementary methods for "Multiomics profiling of human plasma and CSF reveals ATN derived networks and highlights causal links in Alzheimer’s disease"

^q^ Neurology Department, Hospital Sant Pau, Barcelona, Spain. Centro de Investigación en Red en enfermedades neurodegenerativas (CIBERNED).

^r^ University Hospital of Lausanne, Lausanne, Switzerland
^s^ Geriatric Psychiatry, Department of Mental Health and Psychiatry, Geneva university Hospitals, Geneva, Switzerland

^t^ CITA-Alzheimer Foundation, San Sebastian, Spain

^u^ AC Immune SA, Lausanne, Switzerland, formerly Janssen R&D, LLC. Beerse, Belgium at the time of study conduct
^v^ Department of Radiology and Nuclear Medicine, Amsterdam UMC, Vrije Universiteit, The Netherland

^w^ Queen Square Institute of Neurology and Centre for Medical Image Computing, University College London, UK
^x^ Clinical Neurochemistry Laboratory, Sahlgrenska University Hospital, Mölndal, Sweden

^y^ Department of Psychiatry and Neurochemistry, Institute of Neuroscience and Physiology, Sahlgrenska Academy, University of Gothenburg, Mölndal, Sweden
^z^ UK Dementia Research Institute at UCL, London, United Kingdom
^aa^ Department of Neurodegenerative Disease, UCL Institute of Neurology, London, United Kingdom

^bb^ Janssen Medical (UK), High Wycombe, UK

^cc^ Department of Psychology, University of Oslo, Oslo, Norway

**+ contributed equally**

*** Correspondence:**

Cristina Legido Quigley:

Petroula Proitsi:

*Mendelian randomization*

Mendelian randomization (MR) presents a statistical methodology akin to a randomized controlled trial that allows the investigation of putative causal relationships through use of genetic variants as randomizing instruments (1). MR has three key assumptions that must be fulfilled 1) the genetic variants are associated with the exposure of interest (the relevance assumption; 2) that the genetic variants are independent of any confounders (the independence assumption); and 3) that the genetic variants do not influence the outcome of interest, except via the exposure variable (the exclusion restriction assumption).

For AD, we used publicly available summary statistics from the latest GWAS of clinically diagnosed late onset AD by Kunkle et al. (2). For the five circulating hub proteins that that correlated with MCI conversion i.e., PCSK7, RCN2, EFNA2, AP-1 and COL15A1, we used summary statistics from a large proteomics GWA study (3) Data were harmonized between AD and protein datasets, and SNPs with Minor Allele Frequency (MAF) < 0.01 were excluded. All GWAS were assumed to be coded on the forward strand, but sensitivity analyses were performed excluding non-inferable palindromic SNPs (MAF > 0.40).

*Main MR analyses*

For MR analyses with proteins as exposures and AD as an outcome, we identified the lead variants (cis-pQTLs) with the lowest p value for each of the five proteins (3). The Wald ratio method (4) was used to obtain MR effect estimates.

For MR analyses with AD as the exposure and each protein in turn as the outcome, we tested the association of independent Genome Wide Significant (GWS) variants from stages one and two of Kunkle et al. (2). Following clumping (R^2^ threshold = 0.001), removal of variants with MAF<0.01, 20 genetic variants were utilized as instrumental variables in MR analyses. Causal estimates were computed by aggregating variants into a genetic score using IVW two-sample MR.

*Sensitivity analyses*

We applied a number of sensitivity analyses to determine the robustness of the MR findings.

For single cis instrument MR, we first sought to replicate the MR estimates that showed associations using GWA estimates for each protein from an independent Somalogic study (5). Secondly, we used multiple conditional independent cis SNPs to further evaluate the MR findings from our initial MR analysis, since it has been suggested that more precise causal estimates can be obtained using multiple genetic variants from a single gene region, even if the variants are correlated (6, 7). For each protein, we took genetic variants that had a cis association at P_T_ < 5 × 10^−8^ and ‘LD-pruned’ them at r^2^ < 0.3. We then used these genetic variants to construct a genetic score for each protein and used these variants as instrumental variables in univariable IVW MR analyses taking into consideration of their correlation (using the 1000 Genomes European ancestry reference samples), using the Mendelian Randomization R package (8).

For multiple instrument MR analyses two robust methods — MR-Egger (9) and weighted median (10) — were utilized to re-estimate casual associations with IVW assumptions relaxed, as we have previously performed (11) . Additionally, influential points were investigated using leave-one-out analyses, and Cochran’s Q was calculated to test for heterogeneity among instruments (Q—P < 0.05 indicating significant heterogeneity). Finally, due to the strong association of the APOE4 locus with Alzheimer’s disease, we investigated whether results remained the same after removal of variants in the APOE4 locus.

We could not find summary data for the sphingomyelins or phosphatidylethanolamines as A/T/N metabolite hubs associated with MCI conversion status for downstream MR analyses. Analyses were therefore performed using the “Sphingomyelins” NMR metabolite from the Nightingale NMR panel (12).
